# Prediction of the infection of COVID-19 in Bangladesh by classical SIR model

**DOI:** 10.1101/2020.10.21.20216846

**Authors:** Sofi Mahmud Parvez, Faria Tabassum, H. M. Shahadat Ali, Md. Murad Hossain

## Abstract

The ongoing outbreak of the novel coronavirus (COVID-19) started from Wuhan, China, at the end of December 2019. It is one of the leading public health challenges in the world because of high transmissibility. The first patient of COVID-19 was officially reported on March 8, 2020, in Bangladesh. Using the epidemiological data up to October 17, 2020, we try to estimate the infectious size. In this paper, we used Classical SIR (Susceptible-Infected-Recovered), model. The epidemic has now spread to more than 216 countries around the world. The necessary reproduction number *R*_*0*_ of Bangladesh is 1.92. The primary data was collected from the Coronavirus (COVID-19) Dashboard (BANGLADESH: CASE TREND). In our analysis, the statistical parameters specify the best import to provide the predicted result. We projected that the epidemic curve pulling down in Bangladesh will start from the first week of November (November 4, 2020) and may end in the last week of July (July 24, 2021). It is also estimated that the start of acceleration on May 24, 2020, in 53 days, and the start of steady growth on September 10, 2020, in 109 days. The start of the ending phase of the epidemic may appear in the first week of November 2020, and the epidemic is expected to be finished by the last week of July 2021. However, these approximations may become invalid if a large variety of data occurs in upcoming days.

## Introduction

The outbreak of novel coronavirus disease (COVID-19) that was started in Wuhan, China, in December 2019 that has been declared a pandemic. From the world health Organization (WHO), we know that already more than 216 countries have been infected by COVID-19. Globally, about 40 million people have confirmed cases of COVID-19, including more than 1124 thousand deaths as of October 20, 2020 [1]. World Health Organization (WHO) announced the outbreak of coronavirus diseases (COVID-19) after analyzing the one month from reporting of the first case on December 31, 2019in Wuhan, China. The first case was found out in Bangladesh on March 8, 2020 [2]. Among the affected countries highest cases were found in the United States of America (USA), i.e., around8,459,967 patients, by Brazil with 5,251,127 cases and India with 7,602,414.As of October 20, 2020, according to the Worldmeter Covid-19 Data, Bangladesh confirmed 391,586 COVID-19 cases, including 5699 related death cases [3]. In most cases, COVID-19 infection causes only mild illness. However, the disease could be fatal for some elderly people and those with pre-existing medical situations such ashigh blood pressure, diabetes, heart, and respiratory disorders.Respiratory droplets released when someone with the virus sneezes, coughs, or talks, and contact transmission (e.g., touches of contaminated hands to someone’s mouth, nose, or eyes) with an incubation period of 2-14 days are the two most important symptoms of transmission of COVID-19 [4-5]. Almost the virus spread all over the world and outbreak globally. The risk factors for COVID-19 spreading include close contact (e.g., within 2 meters) with an infected person and being coughed or sneezed on by someone who has COVID-19. Since the vaccine of COVID-19 has not yet been fully developed, there is only one to prevent coronavirus practice good hand hygiene,compulsorily using masks and social distancing.Bangladesh declared the first confirmed coronavirus cases after three people tested positive for the infectious virus in the capital Dhaka on March 8, 2020 [6]. Bangladesh first started a nationwide holiday from March 26 to April 4, while the country finds the first coronavirus death on March 18, 2020, ten days after the finding of the first three COVID-19 cases.Bangladesh extended the lockdown until May 30, 2020 [7] and adopted precautionary measures such as a showing of passengers at the airport, suspension of transport, limiting public gathering, including flights, trains, buses, enhancing quarantine duration, devoted COVID-19 hospitals, and enhancing sample test during the lockdown period to halt or to reduce the community spread of this novel deadly virus.In this article, we used a study of the Classical Susceptible-Infected-Recovered (SIR) model [8] for predicting the nature of the spread of COVID-19 in Bangladesh. Using this model, we try to forecasts the epidemic size of Coronavirus in Bangladesh by the quality of data available.

## Methodology

The classical SIR model is an epidemiological model that has been used to predicate the number of infected with a contagious illness in a large population. We divided the population being studied into three different compartments, such as S, I, and R in this method. These three compartments are a function of time t, and they changed by the differential equation system [9,10,11]. S(t) indicates the susceptible individuals. When an infectious individual and susceptible come infectious contact, then susceptible individuals contract the virus and transition to the infectious compartment. I(t) indicates the number of infectious individuals. These are individuals who able to be infected or capable of infecting susceptible individuals. R(t) indicates the number of recovered or deceased individuals. The individuals who have been infected and recovered then entered the remove compartment or died. The number of deaths ignored with respect to the total population (N=S+I+R). For a particular disease in a particular population, these functions may be worked out to estimate possible outbreaks and take them under control. Now we have our differential equations. [12, 13]

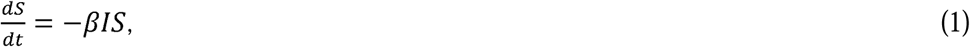

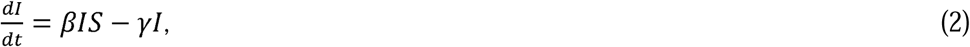

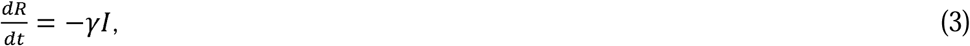

In the above equation, *β* is a positive constant representing a constant rate, and 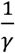 is the average infection period. Particularly these three equations can be reduced to one function about the total infection constant (C=I+R). We collect data from the latest patient case trend data and statistics around coronavirus patient cases for Bangladesh till August 26, 2020 [3]. The simulation of this model was performed using fminsearch, and the ode45 functions of MARAB were implemented according to M. Batista [14].

## Results and Discussion

The outcome of the calculation is presented in Table-1 and in Figure-1. In Table 1, we have found the following calculation*R*_0_(basic reproductive number), *β* (Average interaction frequency), *γ* (average elimination frequency), *C*_{end}_(endemic size), and *S*_{end}_(ending number of susceptible individuals left) with the four episodes of infection, i.e. (*i*) start of acceleration, (*ii*) start of stable growth, (*iii*) start of ending phase and,(*iv*) end of an epidemic (1 case). In Figure-1, we have characterized three graphs: the first one is the total number of novel Corona cases per day, the second is the different epidemic phases, i.e., initial exponential growth, fast growth, asymptotic slow growth, and curve flattening, and the third figure represents daily cases growth factor. The statistical parameters such as coefficient of determination (*R*^*2*^), adjusted *R*^2^, p-value, root mean square error (*RMSE*), and F-statistics vs. zero models for each state are listed in Table 2. The coefficient of determination (*R*^*2*^) and p-value of the model for most of the states are close to 1 and 0, respectively, including the high statistical significance of the results. According to our forecast for Bangladesh from data up to October 17, 2020, the curve flattening of the epidemic will start from the first week of November 2020, and the epidemic may end in the last week of July 2021 (Figure-1). The calculated value of *R*_0_ is 1.153, and the coefficient of determination,*R*^2^=0.773, and p-value (4.65842*e*^-265^) closed to zero indicates the statistical implication of the results found from epidemiological data of Bangladesh. It is worth discussing on the number of samples tested for COVID-19 cases around the world. As of October 17 2020, a total number of 21,92,335 samples have been tested all over the country, of which 391,586 samples were tested positive, which comes out to be 17.8615% of total collected samples (Table-3) [3].

**Figure 1:**
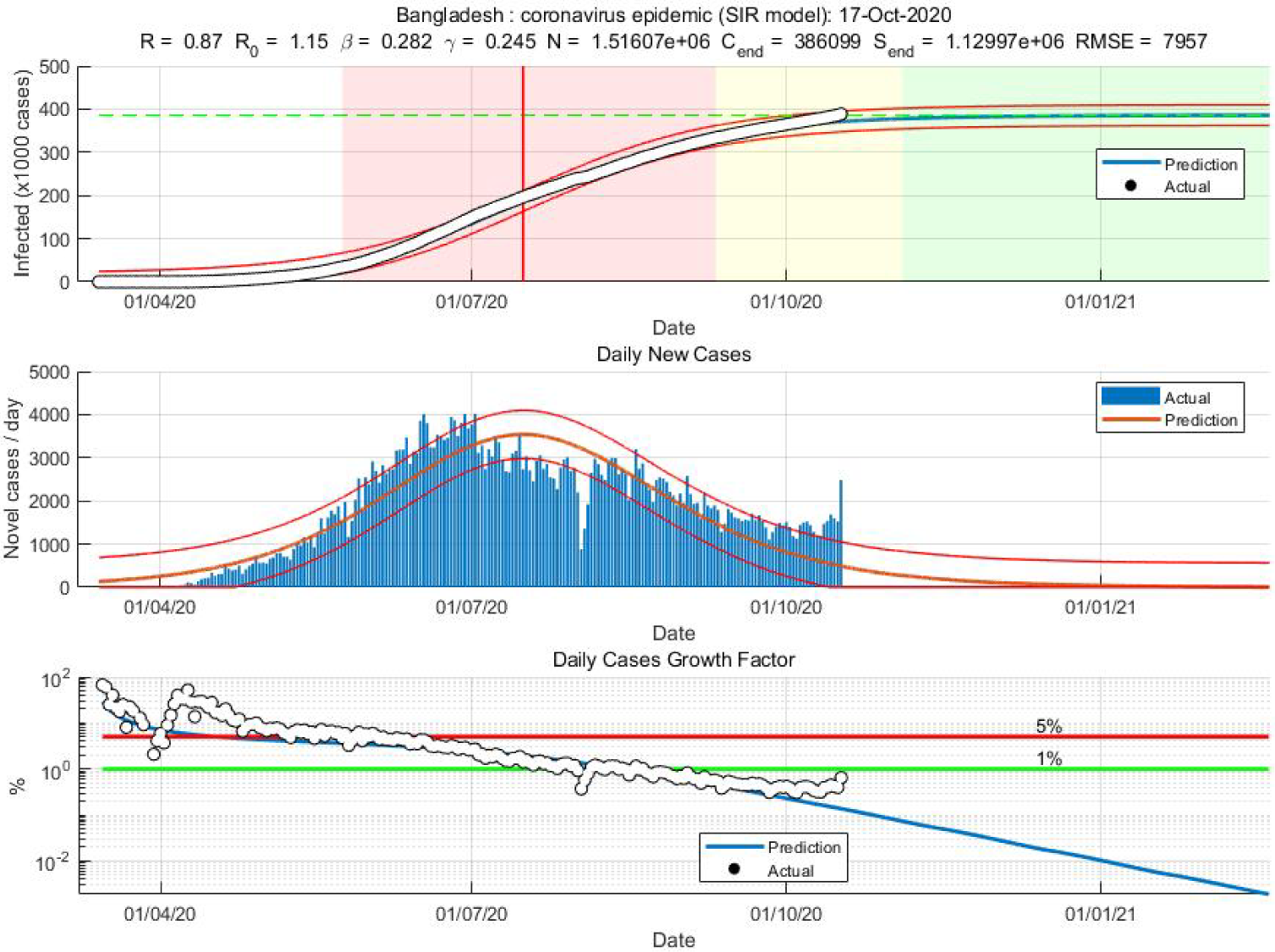
TheForecasts using the SIR model for Bangladesh used data up to October 17, 2020. The individual epidemic phases are highlighted with white, red, yellow, and green colors. Here the upper panel represents initial exponential growth, the middle panel represents fast growth (with positive and negative phase separated by the red vertical line), and the lower panel represents asymptotic slow and curves flatting and daily cases growth factor, respectively.

**Table 1.** Different parameters are calculated by using the SIR model.*R*_0_, *β, γ, C*_{end}_, *S*_{end}_, represent the basic reproductive number, average contact rate, average removal rate, epidemic size, and the final number of susceptible individuals left for Bangladesh.

| $R_0$ | $\beta$ | $\gamma$ | $C_{\{end\}}$ | $S_{\{end\}}$ | Outbreak<br>(year-2020) | Start of<br>acceleration<br>(year-2020) | Start of<br>steady<br>growth<br>(year-2020) | Start of<br>ending<br>phase<br>(year-2020) | End of<br>epidemic<br>(1 case)<br>(year-2021) |
| --- | --- | --- | --- | --- | --- | --- | --- | --- | --- |
| 1.153 | 0.282 | 0.245 | 386099 | $1.12997e^{06}$ | 14-Mar | 24-May | 10-Sep | 4-Nov | 24-July |

**Table 2:** Statisticsof SIR model for Bangladesh

| Number of<br>observations | Degree of<br>freedom | RMSE | $R^2$ | Adjusted R-Squared | F-statistic<br>vs.-zero<br>model | p-value |
| --- | --- | --- | --- | --- | --- | --- |
| 218 | 214 | 7957 | 0.773 | 0.997 | 241489.4 | $4.65842e^{-265}$ |

**Table 3:** Total confirmed cases, Total samples tested, and % of positive cases in Bangladesh as of July 19, 2020 [3]

| Confirmed Cases | Sample tested | % of Positive cases from<br>Tested Samples |
| --- | --- | --- |
| 391,586 | 21,92,335 | 17.8615 |

## Conclusions

In our study, we try to use the classical SIR epidemiological model to forecast the COVID-19 epidemic in the Bangladesh region. The value of the basic reproduction number (RO) is found to be 1.153 for the Bangladesh population. The ending phase of the epidemic may arise in the end week of November 2020, and it may be ended by the last week of July 2021. The forecast is based on the data analysis by MATLAB and that the current deterrent effort will be continued.

## Data Availability

Data will be available upon reasonable request

## References

1. https://www.worldometers.info/coronavirus/COVID-19CORONAVIRUSPANDEMIC

2. Last updated: October 21, 2020, 00:59 GMT

3. WHO Statement on the second meeting of the International Health Regulation (2005). Emergency Committee regarding the outbreak of novel coronavirus (2019-nCoV). Available:https://www.google.com/search?client=firefox-b-d&q=2.%09WHO+Statement+on+the+second+meeting+of+International+Health+Regulations%282005%29+Emergency+Committee+regarding+the+outbreak+of+novel+coronavirus%282019-nCoV%29.2020. Retrieved: 28.08.2020

4. https://www.worldometers.info/Last updated: August 26, 2020.

5. T. Singhal, A review of coronavirus disease-2019 (COVID-19). The Indian Journal of Pediatrics, 2020: p.1–6.

6. J. F. W Chan, S. Yuan, K. K. Hang, H. Chu, and et al., A familial cluster of pneumonia associated with the 2019 novel coronavirus indicating person-to-person transmission: a study of a family cluster. the Lancet, 2020. 395 (10223): p.514–523.

7. https://www.aa.com.tr/en/asia-pacific/bangladesh-confirms-first-case-of-coronavirus-/1758924 Retrieved: March 8, 2020.

8. K. Biswas, A. Khaleque, and P. Sen, Covid-19 spread: Reproduction of data and prediction using a SIR model on Euclidean network, pp. 4–7, 2020, [Online]. Available:http://arxiv.org/abs/2003.07063

9. H. W. Hethcote, The mathematics of infectious diseases. SIAM Review, 2000. 42(4): p.599– 653.

10. D. Guerrero, Spread of Covid-19: a Study Case of Honduras, Forecasting with Logistic Model and SIR Model, Upnfm. Edu. Hn, pp. 1–13, 2020. Available: https://www.upnfm.edu.hn/phocadownload/Covid_19_Danny_Guerrero_Elsarticle.pdf

11. Y. Cha, An Epidemic Model, vol. 6, no. 1, pp. 1–10, 2003. Available:https://www.maplesoft.com/applications/download.aspx?SF=127836/SIRModel.pdf

12. D. Smith and L. Moore, The SIR Model for Spread of Disease - The Differential Equation Model, pp. 1–5, 2020. Available:https://www.maa.org/press/periodicals/loci/joma/the-sir-model-for-spread-of-disease-the-differential-equation-model

13. M. Batista, Estimation of the final size of the COVID-19 epidemic, medRxiv, p. 2020.02.16.20023606, Jan. 2020, DOI: 10.1101/2020.02.16.20023606.

14. M. Batista (2020). fitVirusCOVID19 (https://www.mathworks.com/matlabcentral/fileexchange/74658-fitviruscovid19), MATLAB Central File Exchange). Retrieved May 12, 2020.

